# Longitudinal risk prediction for pediatric glioma with temporal deep learning

**DOI:** 10.1101/2024.06.04.24308434

**Authors:** Divyanshu Tak, Biniam A. Garomsa, Anna Zapaishchykova, Zezhong Ye, Sri Vajapeyam, Maryam Mahootiha, Juan Carlos Climent Pardo, Ceilidh Smith, Ariana M. Familiar, Tafadzwa Chaunzwa, Kevin X. Liu, Sanjay Prabhu, Pratiti Bandopadhayay, Ali Nabavizadeh, Sabine Mueller, Hugo JWL Aerts, Daphne Haas-Kogan, Tina Y. Poussaint, Benjamin H. Kann

## Abstract

Pediatric glioma recurrence can cause morbidity and mortality; however, recurrence pattern and severity are heterogeneous and challenging to predict with established clinical and genomic markers. Resultingly, almost all children undergo frequent, long-term, magnetic resonance (MR) brain surveillance regardless of individual recurrence risk. Deep learning analysis of longitudinal MR may be an effective approach for improving individualized recurrence prediction in gliomas and other cancers but has thus far been infeasible with current frameworks. Here, we propose a self-supervised, deep learning approach to longitudinal medical imaging analysis, temporal learning, that models the spatiotemporal information from a patient’s current and prior brain MRs to predict future recurrence. We apply temporal learning to pediatric glioma surveillance imaging for 715 patients (3,994 scans) from four distinct clinical settings. We find that longitudinal imaging analysis with temporal learning improves recurrence prediction performance by up to 41% compared to traditional approaches, with improvements in performance in both low- and high-grade glioma. We find that recurrence prediction accuracy increases incrementally with the number of historical scans available per patient. Temporal deep learning may enable point-of-care decision-support for pediatric brain tumors and be adaptable more broadly to patients with other cancers and chronic diseases undergoing surveillance imaging.

## INTRODUCTION

Pediatric gliomas, traditionally categorized into high-grade (pHGG) and low-grade gliomas (pLGG) via World Health Organization criteria, originate from glial cells and are the most common type of brain tumors and cause of cancer-related death in children^1^,^2^. Collectively, pLGGs represent a basket of >20 histologies^3^ and, more recently defined, heterogeneous molecular characteristics^4^, with nearly 50% of tumors harboring a BRAF-associated mutation^5^.

Upfront surgical resection, when feasible, is the standard treatment for pediatric gliomas. While pLGGs carry a relatively good prognosis following primary tumor resection compared to high-grade tumors, postoperative outcomes are heterogeneous. Patients undergoing surgical treatment with gross total resection (GTR) typically exhibit favorable outcomes, demonstrating over 85% progression-free survival, yet recurrences still occur^6,7^. Conversely, the prognosis for patients undergoing subtotal resections without subsequent adjuvant therapy is less frequently reported. Available literature indicates that the progression rate ranges from 40% to 80%^8,9^in these cases. The diverse progression patterns inherent in natural history add complexity to decisions regarding adjuvant therapy^10^, a challenge further intensified with the advent of targeted treatments, particularly those for BRAF-associated mutations^5^. In regard to high-grade gliomas, some variations, such as glioblastoma and diffuse midline gliomas, are nearly universally fatal, while others, such as supratentorial grade III tumors, may follow more heterogeneous courses.

Along with histological and molecular subtyping, magnetic resonance imaging (MRI) assessment is critical in determining prognosis and therapy^5^. Early identification of recurrence following surgery optimizes the prospects for salvage interventions to preserve survival and quality of life^10,11^. Despite frequent and long-term MRI surveillance, recurrence can be difficult to assess by diagnosticians given the need to synthesize longitudinal changes over many time points, and, resultingly, recurrence can first present symptomatically^12^.

Computational imaging techniques, particularly deep learning, have demonstrated the ability to synthesize vast quantities of medical imaging data and detect patterns unapparent to the human eye.^13–15^ While there have been several investigations demonstrating the potential for deep learning to extract informative features for pediatric glioma risk stratification, none have leveraged multiple scans as input – *i.e. longitudinal imaging*, which may enable the ability to learn sequential changes in data to better inform risk prediction and decision-making at the point-of-care. Barriers to deep learning-based longitudinal medical imaging analysis include the limited availability of longitudinal datasets and a high dimensional parameter space, which may be unsuitable for supervised deep-learning approaches^16^ due to issues such as curse of dimensionality and parameter explosion. Novel strategies are needed to analyze complex, often long sequences of longitudinal medical imaging data captured clinically. Self-supervised learning (SSL), which utilizes intrinsic information within unlabeled imaging data as a cost-effective supervisory signal for pre-training models,^17–20^ is now viewed as a critical part of medical AI development in limited data scenarios^21^.

Here, we developed an SSL technique, temporal learning, designed specifically for longitudinal medical imaging analysis, and inspired from prior work in self-driving automobile video analysis.^22,23^ We applied temporal learning for the first time to medical imaging to enable longitudinal analysis and risk prediction for pediatric gliomas from serial surveillance scans. We developed and validated the algorithm in multi-institutional setting of longitudinal brain MRIs totaling 3,994 scans from 715 patients with pediatric brain tumors. We found that temporal learning substantially improves recurrence prediction from the time-of-scan for patients with both low- and high-grade gliomas on postoperative surveillance. We expect this study to serve as a foundation for future investigations in longitudinal cancer imaging analyses to enable accurate risk stratification and improved clinical decision-making.

## MAIN

### Postoperative recurrence prediction for pediatric low-grade glioma

#### Datasets

Two retrospective, pediatric, longitudinal, postoperative surveillance MRI datasets with T2 Fluid Attenuated Inversion Recovery (FLAIR) imaging were aggregated for pLGG model development and internal validation: Dana-Farber/Boston Children’s Hospital (DFCI/BCH) pLGG (N=345 patients with low-grade glioma, 2,686 scans, median follow-up: 2,201 days [IQR: 714-3596]) and Children’s Brain Tumor Network (CBTN) (N=301 patients with low-grade glioma, 1,075 scans, median follow-up: 602 [IQR: 247-1457]). A third dataset, RadART (N=35 patients with low-grade glioma, 103 scans, median follow-up: 1,073 days [IQR: 695-3017]), and a fourth dataset, DFCI/BCH pHGG (N=34 patients with high-grade glioma, 130 scans, median follow-up: 154 days [IQR: 89-216]) were curated for external validation of the model in settings of pLGG and pHGG respectively (Figure 2; Methods: Data, Supplementary Methods: A.1, Table S1). All postoperative MRI scans were included up until one year (365 days) prior to last clinical follow-up and/or diagnosis of recurrence, whichever came first. For model development, the DFCI/BCH LGG and CBTN datasets were separately, randomly split into train (80%) and test (20%) sets, with the training set(DFCI/BCH: Npatients=278, Nscans=2175; CBTN: Npatients=240, Nscans=871, respectively) used for model training and validation of the three approaches, and the test set (DFCI/BCH: Npatients=67, Nscans=511; CBTN: Npatients=61, Nscans=204, respectively) reserved as a blinded, holdout set for evaluating recurrence prediction performance of the trained models along with the RadART and DFCI/BCH HGG external sets. We designed a longitudinal imaging deep learning pipeline, comprised of a 3D Resnet18^24,25^ convolutional neural network backbone, a multi-headed self-attention (MHSA), LSTM, and a fully connected layer (Figure 1B, Methods: Deep Learning Pipeline Architecture).

**Figure 1:**
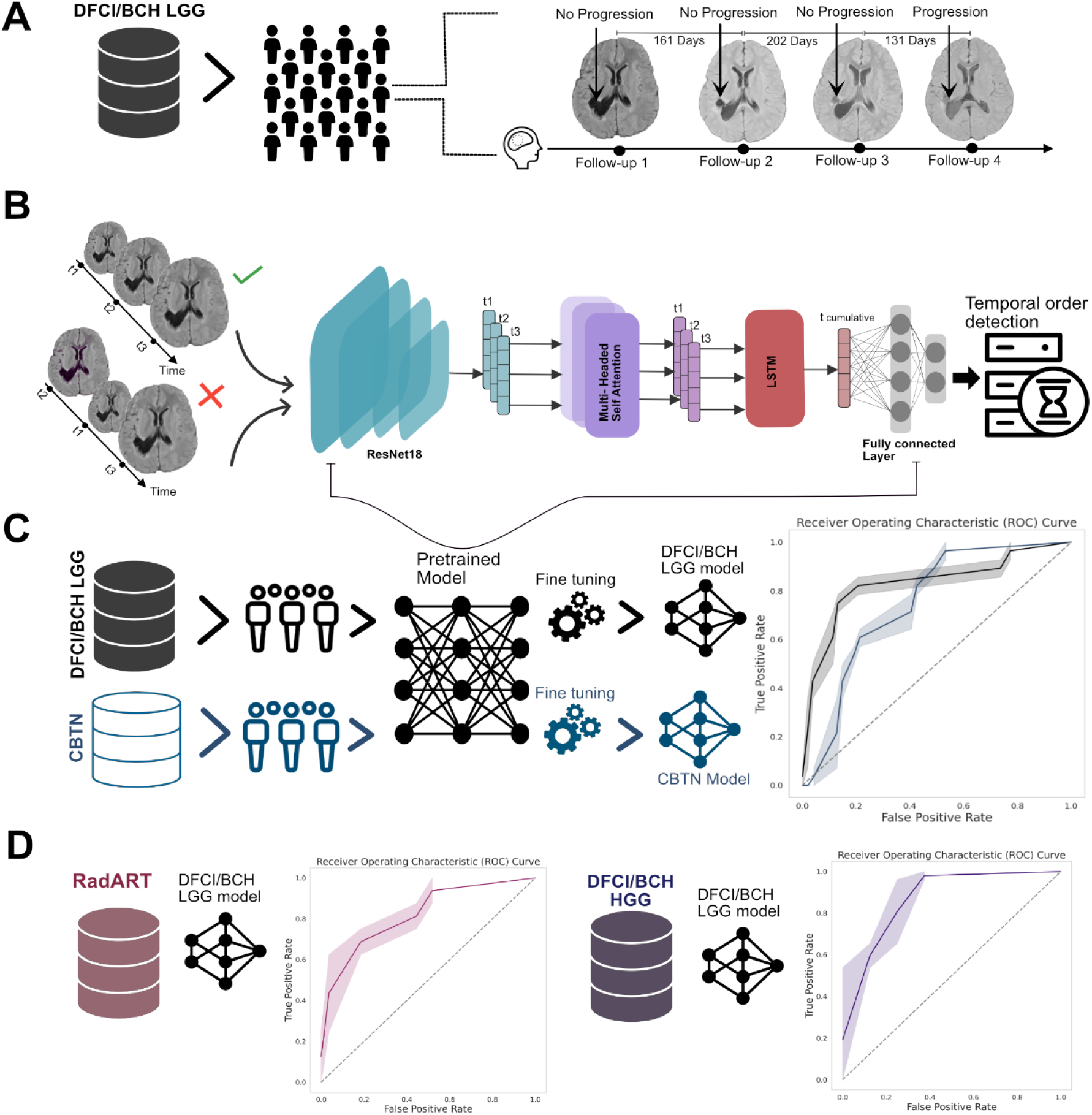
Study design and method overview. A) 4 Datasets with clinical and longitudinal MRI data (T2 FLAIR sequences) were curated from three institutions (n=345 patients, 2686 scans; n=301 patients, 1075 scans; n=35 patients, 103 scans, respectively) for patients with pLGG undergoing postoperative surveillance and (n=34 patients, 130 scans) for patients with pHGG undergoing postoperative surveillance. Each imaging scan was annotated as “progression” or “no progression” based on the radiology report and clinical impression. B) We first train the deep learning pipeline with a self-supervised pretext task, “temporal learning,” to predict whether the input longitudinal imaging sequence is in correct chronological order. The deep learning pipeline consists of 3D ResNet18 encoder followed by multi-headed self-attention block followed by LSTM module appended with a fully connected layer with 2 output neurons for the binary classification. C) We finetune the temporal learning pretrained model to predict the 1-year recurrence risk from time-of-scan. We tested the two fine-tuned models (DFCI/BCH LGG and CBTN) on blinded hold-out sets from each institution (n=67 patients, 511 scans and n=61 patients, 204 scans, respectively). D) We further tested the best-performing finetuned model (DFCI/BCH LGG) on two separate cohorts RadART LGG (n=35 patients, 103 scans) and DFCI/BCH HGG (n=34 patients, 130 scans). Abbreviations: FLAIR=Fluid attenuated inversion recovery, HGG=High grade glioma.

**Figure 2.**
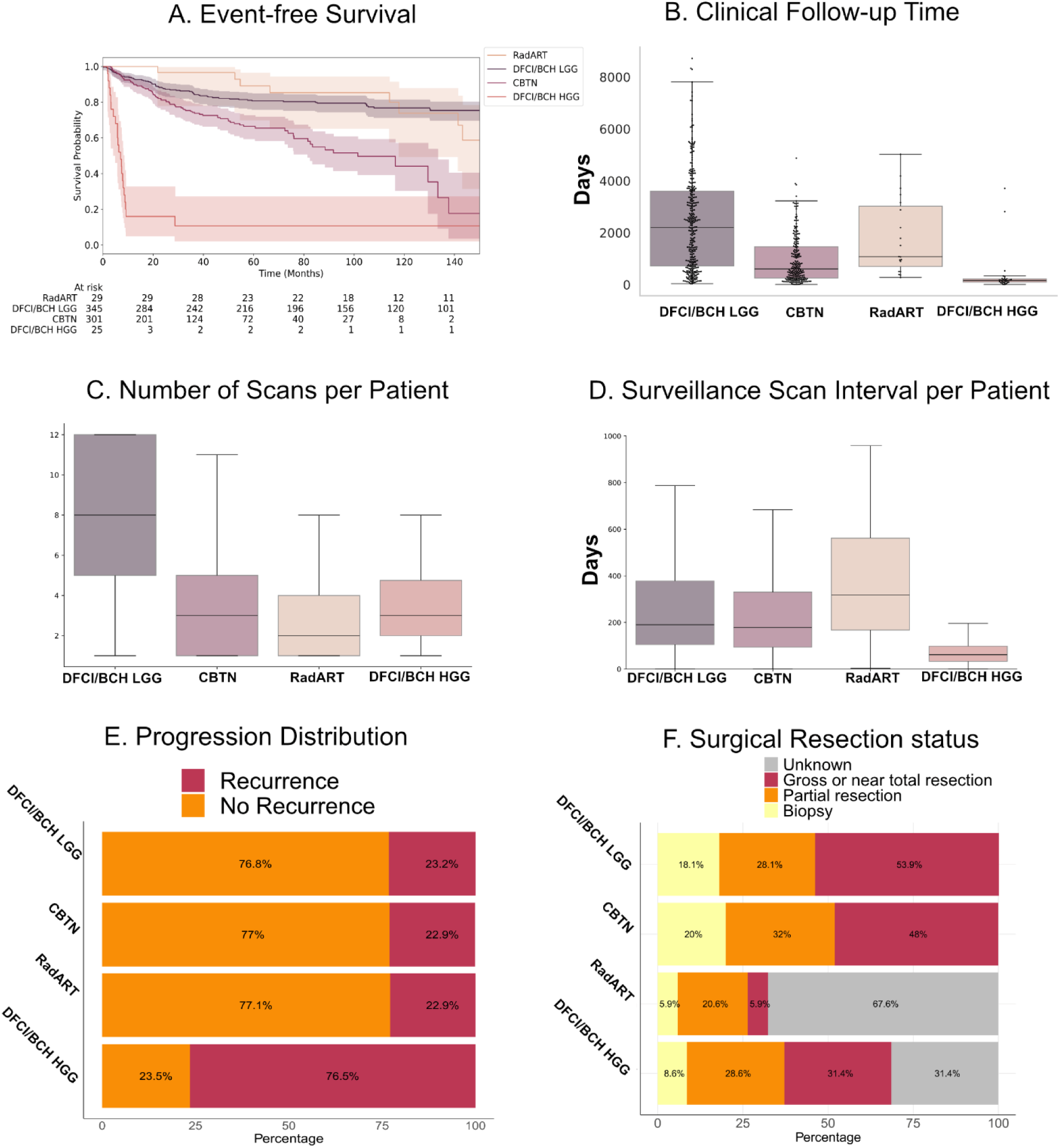
Dataset characteristics. A) Kaplan-Meier event-free survival (EFS) plots for DFCI/BCH LGG (n=345), CBTN (n=301), DFCI/BCH HGG (n=34), and RadART (n=35) cohorts. Patients in the CBTN cohort exhibit a steeper decline in EFS probability over time compared to those in the DFCI/BCH LGG and RadART cohort. Patients in the DFCI/BCH HGG cohort have the steepest decline in survival probability with the worst survival outcomes (log-rank p-value < 0.005) B) Box plot with swarm plot overlay of follow-up time distribution for DFCI/BCH LGG, CBTN, DFCI/BCH HGG and RadART cohorts. The median follow-up times are indicated in the figure: DFCI/BCH LGG with 2201 days [IQR: 714-3596], CBTN with 602 days [IQR: 247-1457], DFCI/BCH HGG with 154 days [IQR: 89-216], and RadART with 1073 days [IQR: 695-3017], as marked by the horizontal black line in each box. The scatter plot overlay represents individual follow-up times, and the whiskers indicate the range excluding outliers. C) Box plot of postsurgical follow-up scan distribution for each cohort before progression. On average, the DFCI/BCH dataset encompasses a greater number of scans per patient when compared to the other cohorts, with the median number of scans per patient being 8 [IQR: 5-12] for the DFCI/BCH, 3 [IQR: 1-5] for the CBTN, 3 [IQR: 2-5] for DFCI/BCH HGG and 2 [IQR: 1-4] for RadART cohort. D) Boxplot of surveillance scan interval distribution per patient. The median surveillance scan interval is 190 days [IQR: 105-378] for DFCI/BCH LGG, 178 [IQR: 94-330] days for CBTN, 61 [IQR: 33-98] days for DFCI/BCH HGG and 317 [IQR: 167-562] days for RadART cohort. E) The bar plot represents the proportion of patients experiencing a recurrence event within the cohorts. F) The bar plot depicts the distribution of surgical resection status among patients in the cohorts. The resection status categories are binned into “Gross or Near total resection”, “Partial Resection”, “Biopsy”, and “unknown”.

The primary endpoint of the study was the prediction of one-year event-free survival (EFS) from the time-of-scan. An event was defined by radiology report impression of recurrence and/or progression, new clinical symptom attributable to tumor, change in patient management, and/or death per retrospective record review. Three approaches to model training were investigated: 1) training from scratch using a single postoperative MRI scan to predict EFS, 2) training from scratch on all longitudinal MRIs to predict EFS, and 3) use of an SSL strategy, temporal learning, for longitudinal MRI model pretraining and then finetuning to predict EFS.

#### Postoperative recurrence prediction with a single scan

We investigated the ability of deep learning to predict one-year EFS based on a single scan input using each postoperative scan for each patient as a model input. The model trained on the DFCI/BCH LGG training set was tested on DFCI/BCH pLGG hold-out set, RadART and DFCI/BCH HGG test sets, while the model trained on CBTN train set was tested on CBTN hold-out set. On the DFCI/BCH LGG test set, the model yielded AUC of 0.58 [95% CI – 0.44-0.73], with sensitivity 0.12, specificity 0.98, and F1 score 0.57 for point-of-scan, one-year EFS prediction. For the CBTN test set, the model yielded AUC of 0.49 [95% CI – 0.44-0.73], with sensitivity 0.21, specificity 0.74, and F1 score 0.46. On the RadART test set the model performed similarly with AUC of 0.43 [95% CI – 0.25-0.67], with sensitivity 0.11, specificity 0.98, and F1 score 0.57. For the DFCI/BCH HGG test set the model yielded AUC of 0.62 [95% CI – 0.52-0.72], with sensitivity 0.61, specificity 0.62, and F1 score 0.61 (Figure 3, Methods: Training Details, Supplementary Methods: A.2).

**Figure 3:**
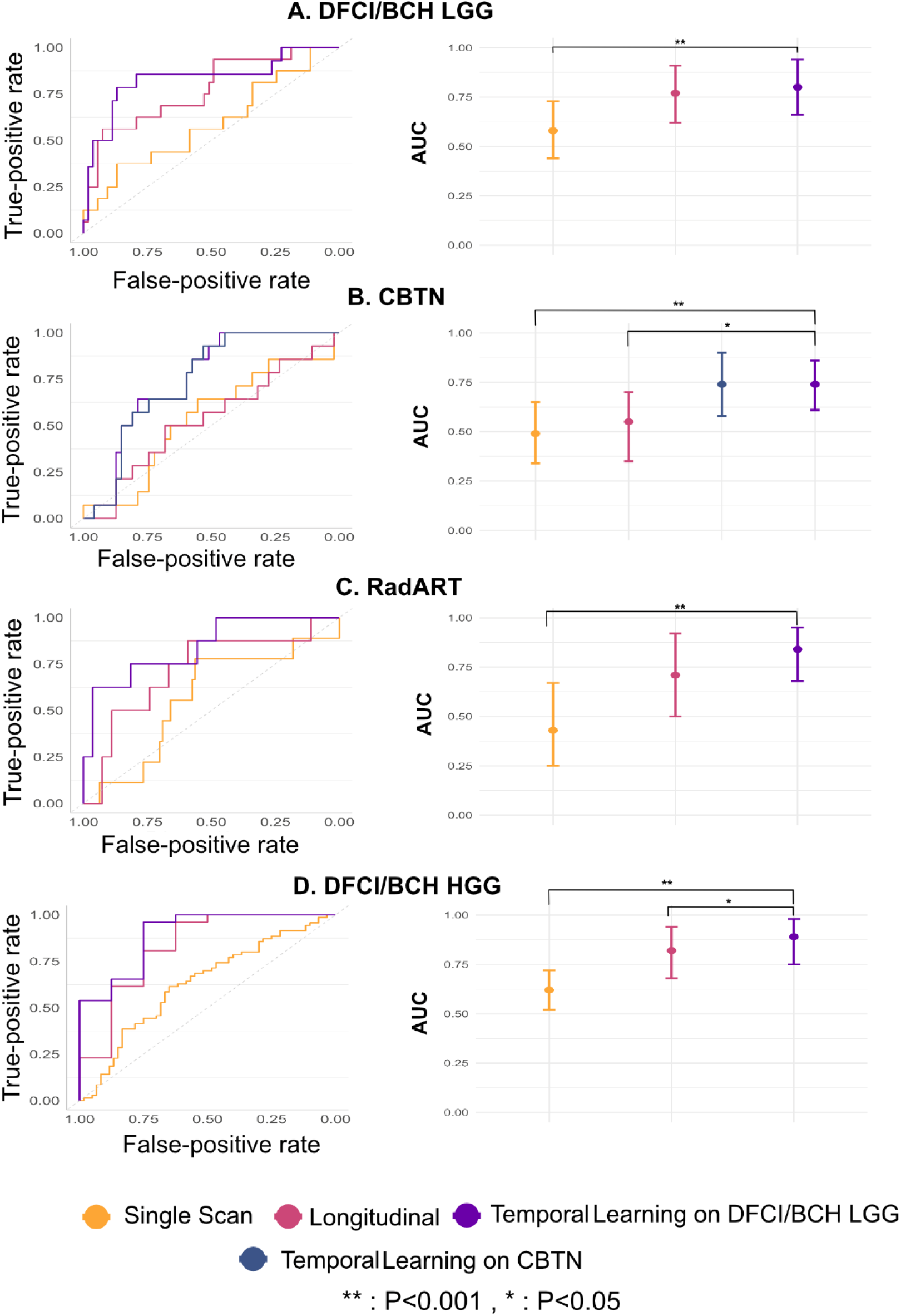
Performance of temporal learning approach to longitudinal imaging analysis, compared with performance of standard longitudinal imaging training and single timepoint imaging approaches. A) ROC curves and AUC comparison plots for the DFCI/BCH LGG dataset. The longitudinal imaging model with temporal learning outperformed the standard longitudinal imaging model and single timepoint imaging model (p=0.003). B) ROC curves and AUC comparison plots for CBTN dataset. The longitudinal imaging model with temporal learning (on DFCI/BCH LGG) performs at par with the longitudinal imaging model with temporal learning on CBTN and outperforms the standard longitudinal imaging model (P=0.01) and single timepoint imaging model (P=0.008) (both on CBTN). C) ROC curves and AUC comparison plots for the RadART dataset, with longitudinal imaging model with temporal learning (on DFCI/BCH LGG) outperforming both standard DFCI/BCH LGG longitudinal imaging model and single timepoint DFCI/BCH LGG imaging model (P=0.0018) on DFCI/BCH LGG cohort. D) ROC curves and AUC comparison plots for the DFCI/BCH HGG dataset. The longitudinal imaging model with temporal learning (on DFCI/BCH LGG) outperforms the standard DFCI/BCH LGG longitudinal imaging model (P=0.03) and single timepoint DFCI/BCH LGG imaging model (P=4.4^e-6^). All p-values are calculated with respect to longitudinal imaging model with temporal learning (on DFCI/BCH LGG), and a two-sided p-value of <0.05 was considered statistically significant.

#### Postoperative recurrence prediction with longitudinal deep learning from scratch

Given the poor performance of single scan prediction, we next investigated patient-level performance using all available longitudinal imaging trained to predict one-year EFS. Sequential scans were passed into the deep learning pipeline using the same training/testing splits, with model trained on DFCI/BCH LGG train set being tested on DFCI/BCH LGG, RadART, DFCI/BCH HGG test set and model trained on CBTN train set tested on CBTN test set. On the DFCI/BCH LGG test set, the longitudinal pipeline yielded an AUC of 0.78 [95% CI – 0.62-0.91], substantially improving over the single timepoint approach (P=0.032), with sensitivity 0.64, specificity 0.77, and F1 score 0.67. On the CBTN test set, AUC was 0.53 [95% CI – 0.35-0.70] with sensitivity 0.50, specificity 0.65, and F1 score 0.55. On the RadART test set the model resulted in the AUC of 0.71 [95% CI – 0.50-0.92] with sensitivity 0.62, specificity 0.70, and F1 score 0.63 (Figure 3A), indicating limited generalizability of learned features across institutions. For the DFCI/BCH HGG test set the model demonstrated improved performance with AUC of 0.82 [95% CI – 0.68-0.94] with sensitivity 0.88, specificity 0.62, and F1 score 0.75 (Figure 3, Table S2).

#### Postoperative recurrence prediction with temporal learning

The temporal learning procedure was designed as follows: a patient’s surveillance scans were input in random chronological sequence and the model was tasked with determining if the sequence was correct (Figure 1B; Methods: Temporal Learning, Supplementary Methods: A.3). Temporal learning was trained and validated by institution, separately, within the DFCI/BCH LGG and CBTN training cohorts.

Additionally, given the significantly longer scan follow-up in the DFCI/BCH LGG dataset, the DFCI/BCH LGG temporal learning trained model was finetuned and tested on the CBTN dataset in parallel to the CBTN temporal learning trained model finetuning, to determine if temporal information learned from one institution with more robust data could generalize to another. Following training procedures, the DFCI/BCH LGG temporal learning model achieved 72% accuracy and area under the receiver operating characteristic curve (AUC) 0.78 [95% CI: 0.64-0.92] in identifying the correct chronological sequence of scans (Supplementary methods A.3). Temporal learning on the CBTN data yielded 66% accuracy and AUC: 0.67 [95% CI – 0.52-0.81].

For one-year EFS prediction, loading the pipeline with weights from temporal learning and finetuning on the DFCI/BCH LGG dataset yielded improved performance on all test sets, including the DFCI/BCH LGG hold-out set (Npatient=67, Nscans=511) with AUC 0.83 [95% CI: 0.71 - 0.91], improved over standard training by 6%, with sensitivity 0.78, specificity 0.86, and F1 score 0.80) (Figure 3, Table S2). The performance improvement on the external CBTN test set was of a larger magnitude with AUC 0.75 [95% CI –0.58- 0.90] and +41.51% increase (P=0.01) and sensitivity 0.71, specificity 0.60, and F1 score 0.65 (Figure 3, Table S2). On the RadART test set the performance improved by 18.32% with AUC 0.84 [95% CI –0.68-0.95], sensitivity 0.62, specificity 0.85, and F1 score 0.73. For the DFCI/BCH HGG test set the model resulted in AUC 0.89 [95% CI – 0.75-0.98] a +8.54% increase with sensitivity 0.88, specificity 0.75, F1 score 0.80. Representative longitudinal cases with predictions are found in Figure 5. Temporal learning on the CBTN dataset also improved EFS prediction compared to training from scratch (Table S2).

**Figure 4.**
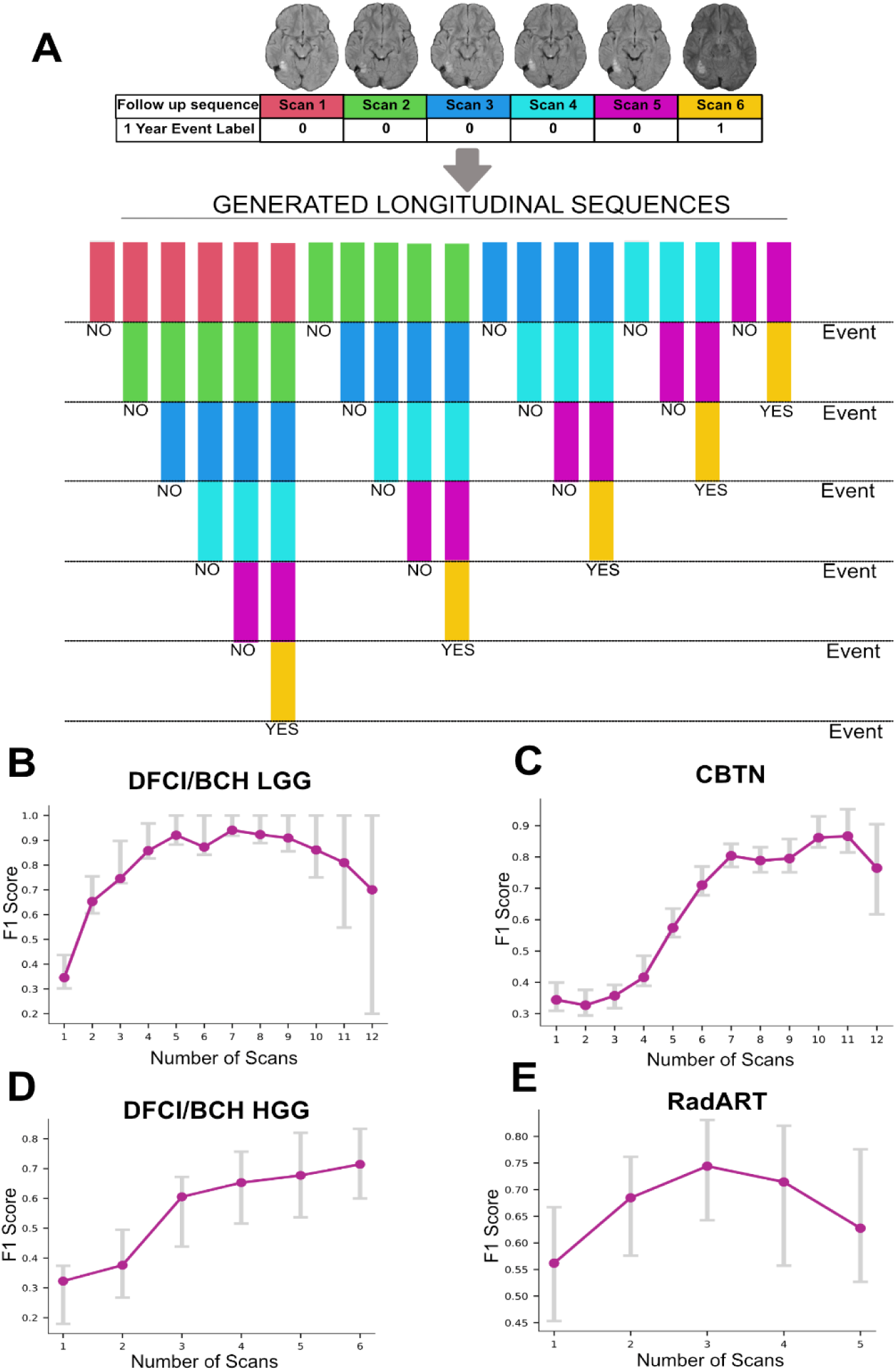
A) Illustration of sequence generation strategy for Intra-patient analysis. The first panel displays the follow-up trajectory of a representative patient with a sequence of follow-up MR scans, and the corresponding binary 1-Year EFS label. The bottom sequences of panels depict the generated trajectories from the original trajectory with the corresponding EFS label. The sequence generation is done iteratively and consecutively starting from the first scan up to the last scan. This technique allows for the estimation of the model’s predictive performance as more longitudinal context is added. B) Intra-patient F1 score plot on DFCI/BCH LGG test set for the model with temporal learning and EFS finetuning on DFCI/BCH LGG train set. The maximum predictive performance is achieved at 7 scans. C) Intra-patient F1 score plot for the CBTN test set for the model with temporal learning on DFCI/BCH LGG train set and EFS finetuning on CBTN train set. The maximum predictive performance is achieved at 10 scans. D) Intra-patient F1 score plot on the DFCI/BCH HGG test set for the model with temporal learning and EFS finetuning on DFCI/BCH LGG train set. The performance increases with increasing number of scans, with maximum performance at 6 scans. E) Intra-patient F1 score plot on the RadART test set for the model with temporal learning and EFS finetuning on DFCI/BCH LGG train set. The maximum predictive performance is reached at 3 scans.

**Figure 5.**
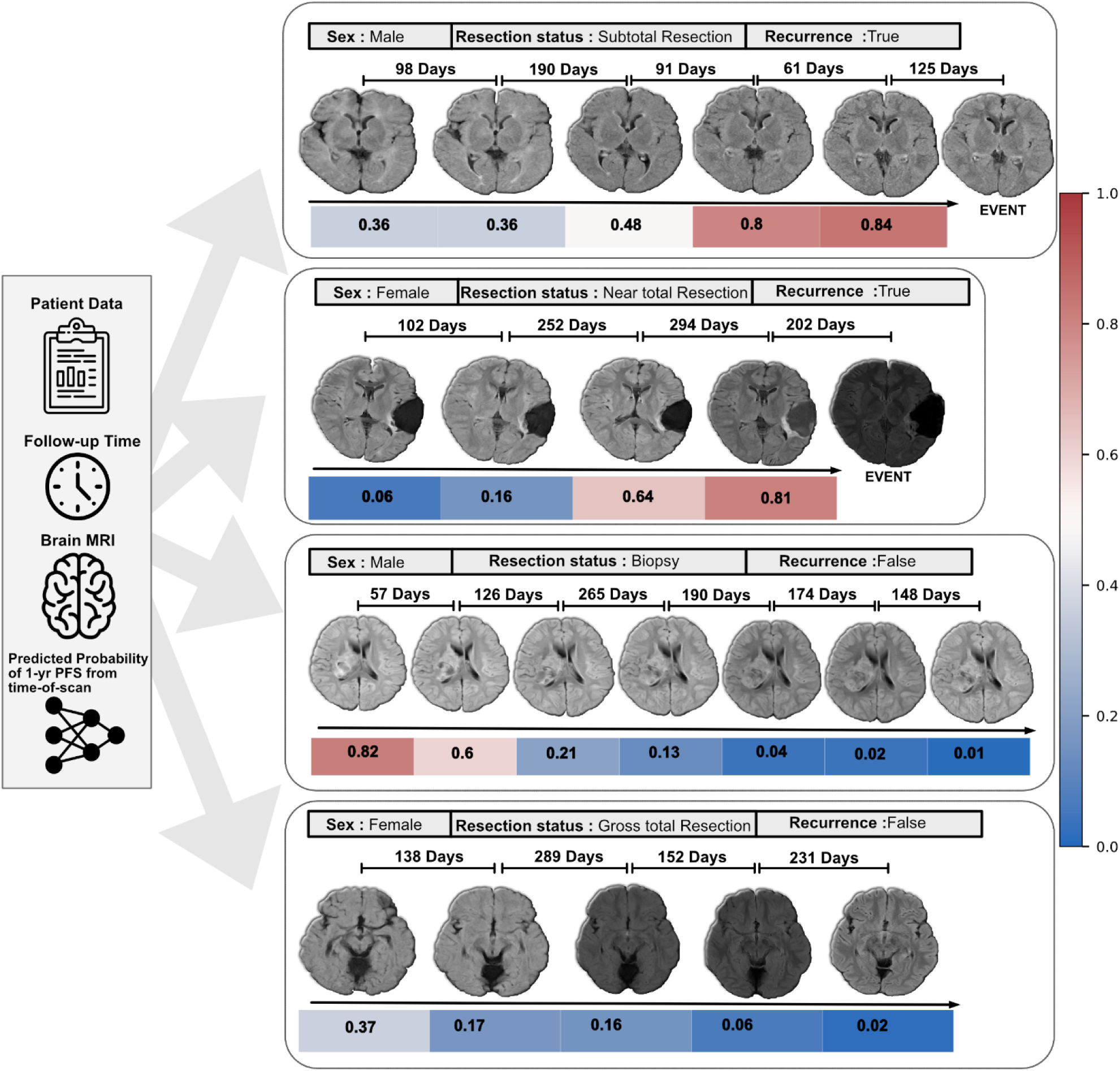
Temporal learning predicts one-year event-free survival in postoperative pLGG patients: representative cases and axial slices from the DFCI/BCH LGG test set. Sequential brain MRI scans demonstrate follow-up intervals and event status in four patients (two males: top and bottom middle; two females: top middle and bottom). Resection status varies among subtotal resection, near total resection, biopsy, and gross total resection. The colored bars indicate the predicted probability output of the pipeline for 1-year event-free survival from time-of-scan by accumulating the postoperative consecutive prior scans up to that point as input, with the corresponding probability of one-year event or death (0.0-1.0) depicted on the right. Event represents radiographic/clinical recurrence or death, and status is marked as either true or false, as the observed clinical outcome for those patients.

### Recurrence prediction performance depends on the number of longitudinal input scans available

To determine how one-year EFS prediction changes with increasing number of input scans, we performed an intra-patient analysis of the DFCI/BCH LGG-trained temporal learning model. For each patient from the test sets of all datasets, we incrementally increased the consecutive longitudinal scans available to the model from one to all and generated one-year EFS predictions from the time of last scan (Figure 4A). Analysis of the DFCI/BCH LGG set revealed the model attained maximum predictive performance (F1 Score) when the number of consecutive input scans reached 7 and then plateaued (Figure 4B). For the CBTN set, the maximum performance reached around 10 scans (Figure 4C), indicating progressive improvements as more longitudinal scans were added. On the DFCI/BCH HGG test set the predictive performance increases with increasing number of scans reaching the maximum at 6 scans (Figure 4D). For the RadART dataset the performance peaks at 3 scans and decreases as further scans are added, though there were very few patients with more than three scans (Figure 4E, Figure S2).

## DISCUSSION

In this work, we leveraged a self-supervised deep learning strategy, temporal learning, for longitudinal MR analysis and postoperative risk assessment in children with gliomas. Up until this point, there have been few attempts to quantitatively analyze longitudinal imaging data for brain tumors^26^, and none for pediatrics, given several barriers, namely limited data and a lack of a framework to synthesize complex, serial imaging data. This study establishes such a framework for longitudinal imaging analysis at the point-of-scan that requires no manual input. We demonstrate that this approach improves the ability to predict postoperative glioma recurrence risk across patients from three institutions and two clinical settings (low- and high-grade glioma), representing over 715 patients and 3994 scans. Deep learning-based short-term risk-stratification may provide an actionable window for early intervention with systemic therapy, radiation, or clinical trial enrollment in patients with high-risk of recurrence. With the advent of novel targeted therapeutics for BRAF-altered gliomas ^27–30^, such a risk-prediction model could inform if, and when, to initiate therapy (Supplementary Methods A.4, Figure S3). Conversely, in those with low projected risk, model outputs may provide reassurance and the ability to de-intensify surveillance regimens.

The cancer care continuum produces a vast amount of longitudinal imaging data containing rich spatiotemporal information and tumor growth patterns. This information may aid in analyzing the response to treatment and predicting recurrence but requires a framework for robustly analyzing potentially billions of parameters of 3D serial medical imaging data without overfitting. We show that training a single timepoint imaging model for the challenging task of postoperative recurrence for pLGG is essentially futile, with model predictions not much better than chance (AUC 0.58 on DFCI/BCH LGG and 0.49 on the CBTN test sets). Adding more longitudinal context in itself was beneficial, but gains in performance disappeared when testing the model on outside data (i.e. CBTN, RadART), demonstrating persistent overfitting and a lack of generalizable features learned. On the other hand, using SSL, as has been widely shown at this point ^31–33^, in helping the model establish a baseline of informative, generalizable features that could be transferred to the setting of recurrence prediction.

Furthermore, our study demonstrates that within an SSL approach, pretext task matters. We hypothesized that the task of learning sequential imaging order (i.e. temporal learning) for postoperative scans with their evolving changes would promote learning of features that inherently capture scan-to-scan differences. Since these differences nearly always occur around residual tumor and surgical cavities, the model inherently learns to focus attention on the clinically relevant areas and assess change from scan-to-scan (Figure 6). This process, in essence, mimics the behavior of an expert neuroradiologist, with the added benefit of the model’s exposure on thousands of sequential scans. The features learned from this process of sequence identification were able to inform accurate recurrence risk prediction with minimal fine-tuning achieving AUCs >=0.75 (Figure 3, Figure S4) across four validation cohorts. Similar strategies have been employed with success in predicting video frame sequences, such as for self-driving cars^22,23^, but had never been investigated for medical imaging. We expect that this strategy will improve medical imaging classification performance across a multitude of scenarios with limited data. While testing of other SSL frameworks is warranted in this setting in the future, pre-existing frameworks are challenging to adapt to multi-timepoint 3D imaging data.

**Figure 6.**
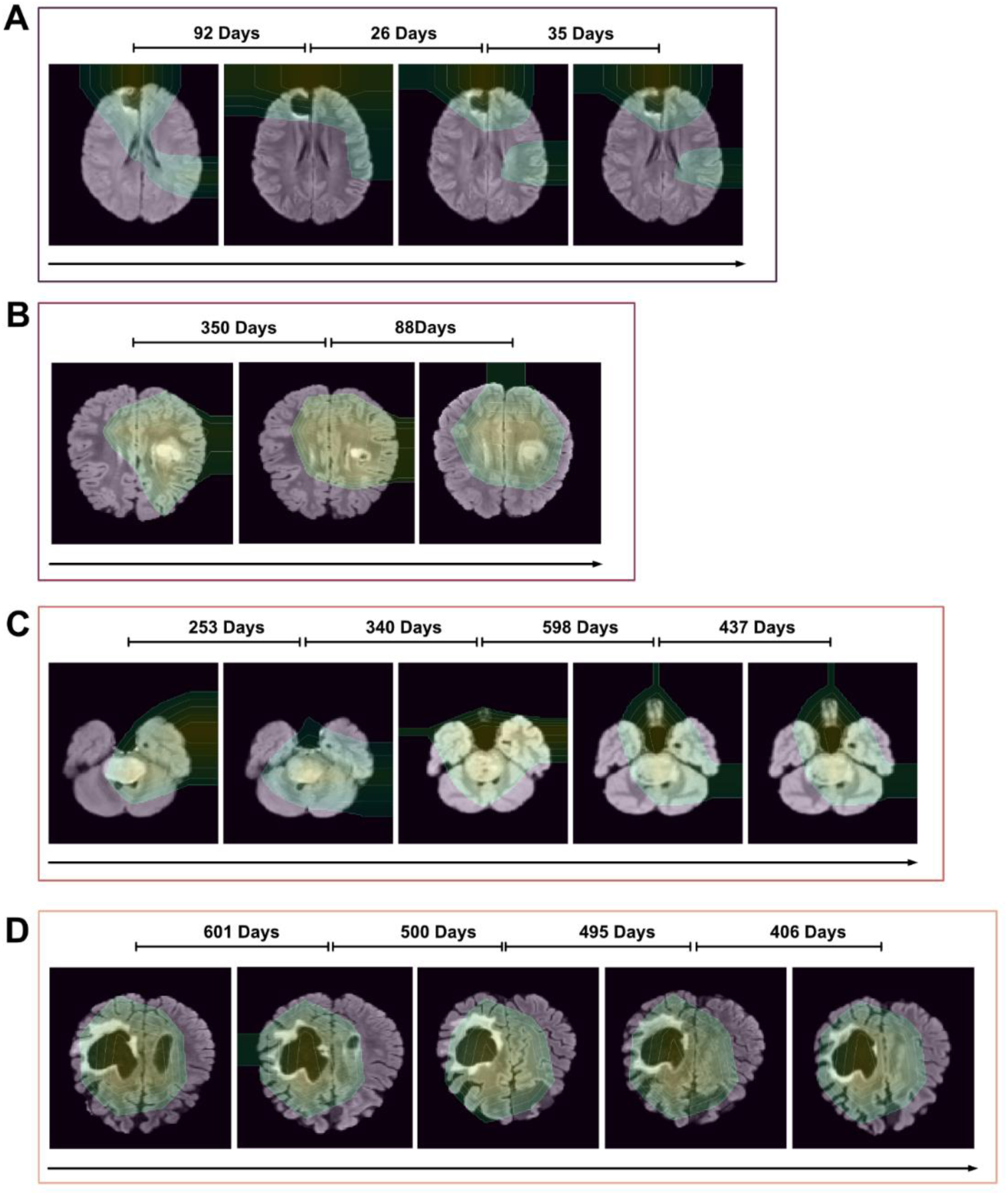
GradCAM maps from the ResNet18 module of the longitudinal imaging pipeline for longitudinal imaging data. The GradCAM map is cutoff at the top 30% intensity with contour lines overlaid. GradCAM visualization for the DFCI/BCH LGG finetuned model (temporal learning on DFCI/BCH LGG) on (A) DFCI/BCH LGG, (C) DFCI/BCH HGG, (D) RadART test subjects. B) GradCAM visualization for the CBTN finetuned model (temporal learning on DFCI/BCH LGG) on CBTN test subject.

To perform highly accurate event predictions, modeling tumor/cavity growth patterns from longitudinal imaging is essential. Previous deep-learning approaches^34–37 38^ have been limited by small datasets and limited longitudinal scans. We address this by leveraging larger datasets with increased longitudinal span. In addition to limited data, prior works lack deeper model designs to capture subtle tumor growth patterns. Our pipeline is designed to pick up inherent spatiotemporal information by encoding spatial cues into latent features using a ResNet encoder, refining features to focus on tumors/cavities with MHSA and modeling temporal dynamics with LSTM. This formed a suitable architecture for temporal learning and, ultimately, recurrence prediction. Notably, temporal learning improved performance regardless of the dataset used for temporal pretraining, though the technique does appear to benefit from pretraining on datasets with longer imaging follow-up (i.e. DFCI/BCH LGG versus CBTN in this study).

This study has several limitations. Firstly, the study is retrospective in nature and subject to selection biases regarding patients included, institutional treatment patterns, surveillance schedules, and variability in MRI scanner acquisition parameters. Prospective validation will be needed to determine the potential effects of data distribution shifts^39^ on performance. There were notably substantial differences in follow-up imaging time and number of scans available across institutions, which may have affected model performance metrics. While incrementally increasing MRI availability was found to be beneficial across institutions, the actual optimal number and point of plateau is likely affected by surveillance intervals, which are not standardized in real-world clinical practice. The scope of this study was limited to one-year EFS prediction, and analysis of longer-term risk prediction would be of clinical utility and a direction for future work. While subgroup analyses by adjuvant therapy were reassuring and showed relative stability (Table S3), further work should investigate the effects of adjuvant therapy on imaging-based recurrence prediction.

In conclusion, we developed and validated the first longitudinal deep-learning imaging model for pediatric brain tumors and found that a self-supervised learning approach inspired by clinical workflows, temporal learning, significantly improved the ability to predict recurrence risk for pediatric gliomas in the postoperative setting and has potential as a point-of-care diagnostic and decision-making tool for cancer surveillance. The temporal learning framework is adaptable to any longitudinal medical imaging task, positioning it for broad impact for disease surveillance and personalized management. Future work is warranted to prospectively validate this framework in a variety of longitudinal imaging scenarios and to study its effects on decision-making and patient outcomes.

## METHODS

### Data

This study was conducted in accordance with the Declaration of Helsinki guidelines and following the approval of local Review Board (IRB). Waiver of consent was obtained from the IRB prior to research initiation due to use of public datasets and retrospective nature of the study. This study involved four patient datasets from three institutions with available T2-weighted-Fluid-Attenuated Inversion Recovery (T2-FLAIR) brain MRI: Dana Farber Cancer Institute/Boston Children’s Hospital (DFCI/BCH) LGG (N=345, median follow-up interval = 190 days) with patients (aged 3 – 20 years) diagnosed between 1990-2022; the Children’s Brain Tumor Network (CBTN; N=301 median follow-up interval = 178 days) with children aged 1 - 23 and pathologically confirmed diagnosis of LGG; RadART (N=35 median follow-up interval = 317 days) with patients aged 5 – 24 years, and diagnosed with LGG; DFCI/BCH HGG (N=34 median follow-up time interval = 61 days) with patients (aged 3-19) diagnosed with HGG. All datasets include T2-FLAIR MRI of patients who underwent primary surgery and had at least one year of clinical and radiographic follow-up. T2-FLAIR MRI images pose superior sensitivity in detecting white matter abnormalities and lesions, along with suppressing CSF and enhancing the contrast between normal and pathological tissues, hence play a crucial role in monitoring tumors and assessing treatment efficacy. These properties also allow deep-learning models to learn better representations of tumor regions for tasks like classification and segmentation. We develop our pipeline around the T2-FLAIR MR sequences for these advantages. MR acquisition details for both datasets are provided in supplemental material (Table S1, Figure S1; Supplementary methods A.1).

### Deep Learning Pipeline Architecture

The single scan imaging model consists of a 3D ResNet18 CNN backbone up to the global average pool layer, followed by a fully connected layer of 512 neurons with 2 output neurons. The longitudinal deep-learning pipeline consists of a 3D ResNet18 CNN backbone up to the global average pool layer. The backbone is followed by a Multi-Headed Self Attention MHSA block with 8 attention heads that accept the latent features of dimension 512. The MHSA block is followed by an LSTM module with the same feature and hidden state dimension of 512. The LSTM module is followed by a Fully Connected layer with 512 neurons and the output layer with 2 neurons (Figure 1B).

### Temporal Learning

we leverage the data from the training split of the DFCI/BCH LGG cohort to prevent information leakage. To implement temporal pretraining, we sample each patient trajectory to create new data points with different scan lengths ranging from 1 to the complete length of the trajectory. We then shuffle all the trajectories and assign labels based on the correct chronological order of the scans for each shuffled trajectory help(0 for incorrect order, and 1 for correct order) (Figure 7). Through this oversampling process, we create 3531 new trajectories from 278 patient trajectories (DFCI/BCH LGG), with number of scans ranging up to 12 per trajectory. We then train and validate the model on this new oversampled dataset and use the best checkpoint for the finetuning process for the EFS task on both the DFCI/BCH LGG and CBTN datasets (Supplementary methods A.3). We perform similar oversampling strategy and temporal learning experiments on the CBTN train split (2054 trajectories from 240 patient trajectories), and then choose the best performing temporal learning model and fine-tune it for EFS prediction task on the CBTN dataset. We compare the EFS predictive performance of the finetuned model with temporal learning on DFCI/BCH LGG and CBTN cohorts (Figure 3, Table S2).

**Figure 7.**
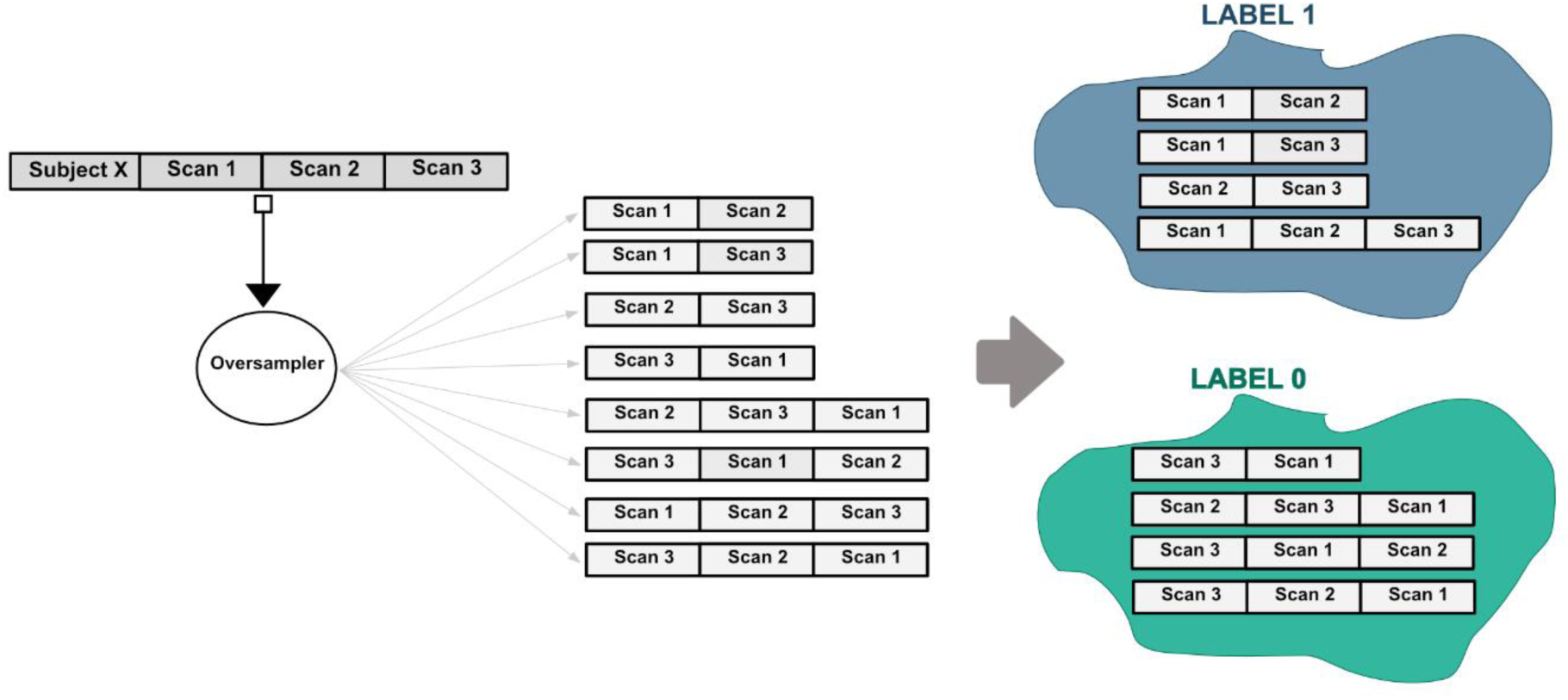
Illustration of oversampling strategy for Temporal Training. The left panel depicts a subject with 3 follow-up scans in chronological order. The middle panel depicts the generated trajectories for temporal learning. Oversampling is done such that the generated trajectories have a variable number of scans starting from 1 up to the maximum scans for that subject. The trajectories are shuffled to generate both positive samples (correct chronological order) and negative samples (incorrect chronological order). Finally, each trajectory is assigned a label (0,1) based on the chronological order of the scans.

### Training details

#### Dataset

T2-FLAIR MRI images from the 4 datasets (Figure 2) were converted from DICOM format to NIFTI format via rasterization packages utilizing dcm2nii package in Python v3.8. N4 bias filed correction was adopted to correct the low-frequency intensity non-uniformity present in MRI images using SimpleITK in Python v3.8. All scans were resampled to 1′1′1 mm^3^ voxel size using linear interpolation via SimpleITK. After interpolation, the MRI scans were co-registered using a rigid registration step with SimpleITK. Lastly, a brain extraction step was performed for all the scans using HD-BET package with final image dimensions of (170,206,162).

For each follow-up scan of a patient, we create a binary 1-year EFS label, based on the difference between the date of the scan and the date of the event. The label is 1 if the difference is less than 365 days and 0 otherwise. In this way, each follow-up scan in the patient trajectory is assigned a binary label with the last scan of the trajectory indicating either the occurrence of the event (label 1) or no event (label 0) for that patient. A split of 80:20 was used for training and validation for training from scratch, temporal pretraining, and finetuning for both DFCI/BCH LGG and CBTN datasets.

#### Dataloader

As the first preprocessing step, we perform intensity standardization across the scans for each patient. This is followed by data augmentations of Random Affine transformation, 3D Elastic transformation, Random Adjust Contrast transformation, Random Rotate transformation, and Intensity scaling using MONAI API. The images were finally resized to a dimension of (96,128,96) before being fed into the model.

#### Training

For the model training, validation, and testing (Supplementary methods A.2), the maximum number of scans per patient was limited to 12. For each patient, all the scans post augmentations are collated into a single list of length 12. For patients with scans less than 12, the scans are appended with empty arrays of the same dimensions to fill the list, whereas for the patients with scans more than 12, the scans are subsampled from the trajectory, keeping the first and the last scans constant.

#### Evaluation

For Training, testing, and reporting of results, we calculate the AUC with 95% confidence intervals, F1 score, specificity, and sensitivity. The 95% confidence intervals are calculated by the bootstrapping method with 1000 bootstrap samples. Statistical metrics and curves were calculated using Scikit-learn packages^40^ in Python v3.8.

## List of abbreviations

pLGG: pediatric Low-Grade Glioma
CNN: Convolutional neural network
AUC: Area under the curve
DL: Deep learning
MRI: Magnetic resonance imaging
SSL: Self-supervised Learning
MHSA: Multi-headed self-attention
LSTM: Long short-term memory network
FC: Fully connected layer
HGG: High-grade glioma
AI: Artificial Intelligence

## DATA AVAILABILITY

All data supporting the findings described in this manuscript are available in the article, in the Supplementary Information, and from the corresponding author upon request. The CBTN data is available upon request at “ https://cbtn.org/ “. The DFCI/BCH and RadART brain tumor dataset contains private hospital data that is controlled due to privacy concerns. Access to the derived dataset will be considered upon request to the corresponding author (Benjamin H. Kann, M.D., timeframe for response 2 weeks).

## Acknowledgments

This study was supported in part by the National Institute of Health/ the National Cancer Institute (NIH/NCI) (U54 CA274516 and P50 CA165962), and Botha-Chan Low Grade Glioma Consortium. We would also like to acknowledge the Children’s Brain Tumor Network (CBTN) for the imaging and clinical data access.

## Author Contributions

Study design: D.T., and B.H.K.; code design, implementation and execution: D.T.; acquisition, analysis or interpretation of data: D.T., A.Z., and B.H.K.; writing of the manuscript: D.T., B.H.K.; critical revision of the manuscript for important intellectual content: all authors; statistical analysis: D.T.; study supervision: B.H.K., H.J.W.L.A., T.P., and D.H.K.

## CODE AVAILABILITY

The model source code will be made available at time of publication via Github repository.

## REFERENCES

1. Pollack, I. F. Brain Tumors in Children. New England Journal of Medicine 331, 1500–1507 (1994).

2. Packer, R. J. et al. Pediatric low-grade gliomas: implications of the biologic era. Neuro Oncol 19, 750–761 (2017).

3. Manoharan, N., Liu, K. X., Mueller, S., Haas-Kogan, D. A. & Bandopadhayay, P. Pediatric low-grade glioma: Targeted therapeutics and clinical trials in the molecular era. Neoplasia 36, 100857 (2023).

4. Ryall, S., Tabori, U. & Hawkins, C. Pediatric low-grade glioma in the era of molecular diagnostics. Acta Neuropathologica Communications 8, 30 (2020).

5. Dodgshun, A. J., Hansford, J. R. & Sullivan, M. J. Risk assessment in paediatric glioma—Time to move on from the binary classification. Critical Reviews in Oncology/Hematology 111, 52–59 (2017).

6. Bandopadhayay, P. et al. Long-Term Outcome of 4,040 Children Diagnosed With Pediatric Low-Grade Gliomas: An Analysis of the Surveillance Epidemiology and End Results (SEER) Database. Pediatr Blood Cancer 61, 1173–1179 (2014).

7. Wisoff, J. H. et al. Primary Neurosurgery for Pediatric Low-Grade Gliomas: A Prospective Multi-Institutional Study From the Children’s Oncology Group. Neurosurgery 68, 1548 (2011).

8. Gnekow, A. K. et al. Long-term follow-up of the multicenter, multidisciplinary treatment study HIT-LGG-1996 for low-grade glioma in children and adolescents of the German Speaking Society of Pediatric Oncology and Hematology. Neuro-Oncology 14, 1265–1284 (2012).

9. Stokland, T. et al. A multivariate analysis of factors determining tumor progression in childhood low-grade glioma: a population-based cohort study (CCLG CNS9702). Neuro-Oncology 12, 1257–1268 (2010).

10. Collins, K. L. & Pollack, I. F. Pediatric Low-Grade Gliomas. Cancers (Basel) 12, 1152 (2020).

11. de Blank, P., Bandopadhayay, P., Haas-Kogan, D., Fouladi, M. & Fangusaro, J. Management of Pediatric Low-Grade Glioma. Curr Opin Pediatr 31, 21–27 (2019).

12. Malik, D. G. et al. Advanced MRI Protocols to Discriminate Glioma From Treatment Effects: State of the Art and Future Directions. Front. Radiol. 2, (2022).

13. Automated temporalis muscle quantification and growth charts for children through adulthood | Nature Communications. https://www.nature.com/articles/s41467-023-42501-1.

14. Kann, B. H. et al. Screening for extranodal extension in HPV-associated oropharyngeal carcinoma: evaluation of a CT-based deep learning algorithm in patient data from a multicentre, randomised de-escalation trial. The Lancet Digital Health 5, e360–e369 (2023).

15. Cao, K. et al. Large-scale pancreatic cancer detection via non-contrast CT and deep learning. Nat Med 29, 3033–3043 (2023).

16. Deep Learning. https://www.deeplearningbook.org/.

17. Huang, Z., Bianchi, F., Yuksekgonul, M., Montine, T. J. & Zou, J. A visual–language foundation model for pathology image analysis using medical Twitter. Nat Med 29, 2307–2316 (2023).

18. Moor, M. et al. Foundation models for generalist medical artificial intelligence. Nature 616, 259–265 (2023).

19. Zhou, Y. et al. A foundation model for generalizable disease detection from retinal images. Nature 622, 156–163 (2023).

20. Towards a general-purpose foundation model for computational pathology | Nature Medicine. https://www.nature.com/articles/s41591-024-02857-3.

21. Tak, D. et al. Noninvasive Molecular Subtyping of Pediatric Low-Grade Glioma with Self-Supervised Transfer Learning. Radiology: Artificial Intelligence e230333 (2024) doi:10.1148/ryai.230333.

22. Xu, D. et al. Self-Supervised Spatiotemporal Learning via Video Clip Order Prediction. in 2019 IEEE/CVF Conference on Computer Vision and Pattern Recognition (CVPR) 10326–10335 (IEEE, Long Beach, CA, USA, 2019). doi:10.1109/CVPR.2019.01058.

23. Lang, C., Braun, A., Schillingmann, L., Haug, K. & Valada, A. Self-Supervised Representation Learning from Temporal Ordering of Automated Driving Sequences. Preprint at http://arxiv.org/abs/2302.09043 (2023).

24. Papers with Code - ResNet 3D. https://paperswithcode.com/lib/torchvision/resnet-3d.

25. Tran, D. et al. A Closer Look at Spatiotemporal Convolutions for Action Recognition. Preprint at http://arxiv.org/abs/1711.11248 (2018).

26. Lee, C. et al. Applying artificial intelligence to longitudinal imaging analysis of vestibular schwannoma following radiosurgery. Sci Rep 11, 3106 (2021).

27. Talloa, D. et al. BRAF and MEK Targeted Therapies in Pediatric Central Nervous System Tumors. Cancers (Basel) 14, 4264 (2022).

28. Manoharan, N., Liu, K. X., Mueller, S., Haas-Kogan, D. A. & Bandopadhayay, P. Pediatric low-grade glioma: Targeted therapeutics and clinical trials in the molecular era. Neoplasia 36, 100857 (2022).

29. Kilburn, L. B. et al. The type II RAF inhibitor tovorafenib in relapsed/refractory pediatric low-grade glioma: the phase 2 FIREFLY-1 trial. Nat Med 30, 207–217 (2024).

30. Sidaway, P. Tovorafenib effective against low-grade gliomas harbouring BRAF fusions. Nat Rev Clin Oncol 21, 83–83 (2024).

31. Pai, S. et al. Foundation model for cancer imaging biomarkers. Nat Mach Intell 6, 354–367 (2024).

32. Azizi, S. et al. Robust and data-efficient generalization of self-supervised machine learning for diagnostic imaging. Nat. Biomed. Eng 7, 756–779 (2023).

33. Krishnan, R., Rajpurkar, P. & Topol, E. J. Self-supervised learning in medicine and healthcare. Nat. Biomed. Eng 6, 1346–1352 (2022).

34. Lee, G., Kang, B., Nho, K., Sohn, K.-A. & Kim, D. MildInt: Deep Learning-Based Multimodal Longitudinal Data Integration Framework. Frontiers in Genetics 10, (2019).

35. Zhang, L. et al. Spatio-Temporal Convolutional LSTMs for Tumor Growth Prediction by Learning 4D Longitudinal Patient Data. Preprint at http://arxiv.org/abs/1902.08716 (2019).

36. Wang, C. et al. Toward predicting the evolution of lung tumors during radiotherapy observed on a longitudinal MR imaging study via a deep learning algorithm. Medical Physics 46, 4699–4707 (2019).

37. Jin, C. et al. Predicting treatment response from longitudinal images using multi-task deep learning. Nat Commun 12, 1851 (2021).

38. Lu, L., Dercle, L., Zhao, B. & Schwartz, L. H. Deep learning for the prediction of early on-treatment response in metastatic colorectal cancer from serial medical imaging. Nat Commun 12, 6654 (2021).

39. Finlayson Samuel G., et al. The Clinician and Dataset Shift in Artificial Intelligence. New England Journal of Medicine 385, 283–286 (2021).

40. scikit-learn: machine learning in Python — scikit-learn 1.4.0 documentation. https://scikit-learn.org/stable/.

