## Supplementary for "Longitudinal risk prediction for pediatric glioma with temporal deep learning"

### Supplementary Methods

#### A.1 Dataset acquisition details

Patients from the Children's Brain Tumor Network (CBTN) cohort underwent brain MR imaging at 1.5T or 3T across various vendors (Siemens; GE Healthcare; Philips Healthcare; Hitachi; and Toshiba). Because the CBTN imaging data was collected across several institutions as part of the clinical standard of care, images were acquired with non-uniform acquisition protocols. Patients from the DFCI/BCH underwent brain MR imaging at 1.5T or 3T from various MRI vendors (Siemens; GE Healthcare). Patients from the RadART cohort underwent brain MR imaging from vendors (GE, Siemens, Phillips) at 1.5T or 3T. All MR imaging data were extracted from the respective PACS and metadata were de-identified for analyses.

#### A.2 Deep-Learning Training details

The single timepoint imaging model consists of a 3D ResNet18 encoder up to the global average pool layer. The fully connected layer from the native ResNet18 was replaced by a fully connected layer of 512 neurons followed by an output layer of 2 neurons. The longitudinal imaging model builds on top of the single timepoint imaging model by the addition of a Multi-Headed Self Attention (MHSA) block (feature dimension 512, and the number of self-attention heads = 8) and an LSTM (feature dimension 512, and latent dimension 512) block before the fully connected layer of 512 neurons which is followed by the output layer of 2 neurons.

The models were trained for a binary classification task of 1Year EFS for 250 epochs with a batch size of 8, Stochastic Gradient Descent SGD optimizer, a learning rate of 0.0005, momentum of 0.9, and a weight decay of 0.00005. A cyclic learning rate scheduler was used with a step size of 2000 and triangular mode. The development, training, and validation of the pipeline was done in Python 3.8 with PyTorch deep-learning framework (Version 2.0.1 + Cuda 11.7) on Nvidia A6000 GPU. The 3D implementation of ResNet18 was used from the MONAI framework, and MHSA and LSTM modules were imported from PyTorch.

The preprocessed MRI scans with image dimensions of (96,128,96) were collated into a list of length 12 for each patient, with appending empty arrays of the same dimensions for patients having less than 12 scans. Random Affine transformation, 3D Elastic transformation, Random Adjust Contrast transformation, Random Rotate transformation, and Intensity scaling were applied to the collated list with the same transformation parameters for each scan in the list. The transformations were imported from MONAI API.

#### A.3 Temporal Learning

Temporal learning was performed on the training split of the DFCI/BCH LGG (278 patients and 2175 scans) and CBTN (240 patients and 871 scans) datasets. After oversampling the DFCI/BCH LGG set resulted in 3531 trajectories, and CBTN set resulted in 2054 trajectories. With the training time and memory constraints, a generation of a maximum of 20 samples was limited from a single trajectory. Each generated trajectory was labeled based on the chronological order of the scans (0 for incorrect chronological order and 1 for correct chronological order). A random split of 80:20 was used for training and validation of the temporal learning model. The best performing temporal learning model on DFCI/BCH LGG results in AUC of 0.78 [95% CI – 0.64-0.92], sensitivity 0.78, specificity 0.70, and F1-Score 0.74. While the best performing temporal learning model on CBTN could classify between the correct and incorrect chronological order sequences with AUC 0.67 [95% CI – 0.52-0.81], sensitivity 0.65, specificity 0.68, and F1-Score 0.67.

#### A.4 EFS prediction performance in BRAF-mutated gliomas

We analyzed the performance of the temporal pretraining model finetuned on DFCI/BCH LGG and CBTN cohort separately, on the subgroup of patients with a pathologically confirmed BRAF-mutation (V600E point-mutation, fusion, or wildtype) (Figure S5). For the DFCI/BCH test the model results in the AUC 0.74 [95% CI – 0.60-0.91], sensitivity 0.75, specificity 0.88, and F1 Score 0.75. For the CBTN test the model results in AUC 0.80 [95% CI – 0.64-0.92], sensitivity 0.60, specificity 0.78, F1 Score 0.66.

#### Supplementary Figures

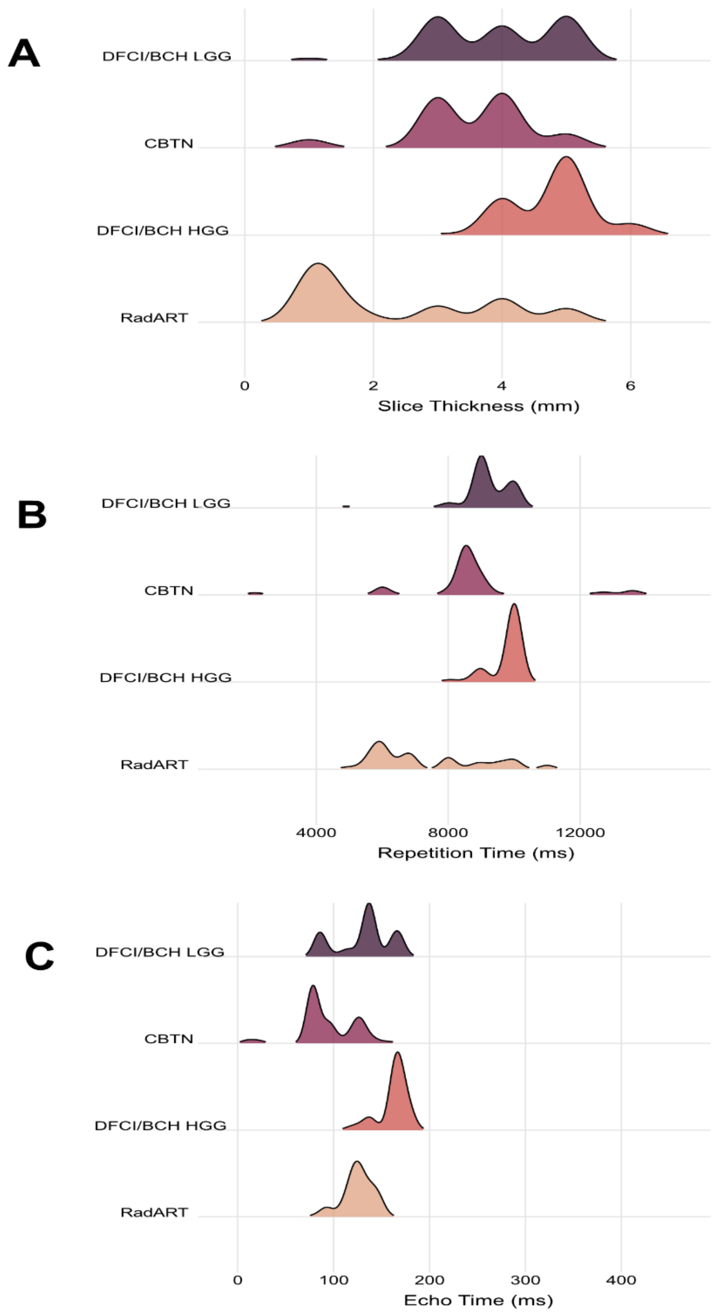

Figure S1. MR Acquisition distribution comparison for DFCI/BCH LGG, CBTN, RadART, and DFCI/BCH HGG Datasets for A) Slice thickness, B) Repetition Time, and C) Echo Time.

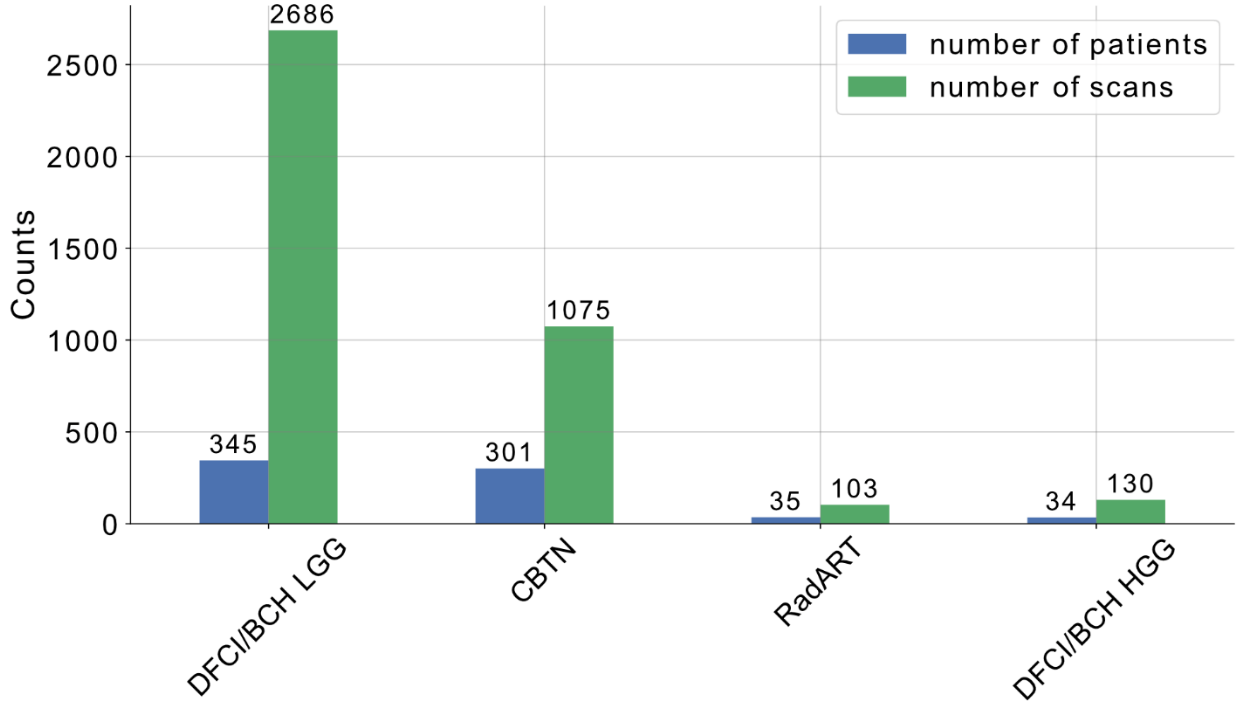

Figure S2. Patient and Scan Distribution for DFCI/BCH LGG, CBTN, RadART, and DFCI/BCH HGG Cohort.

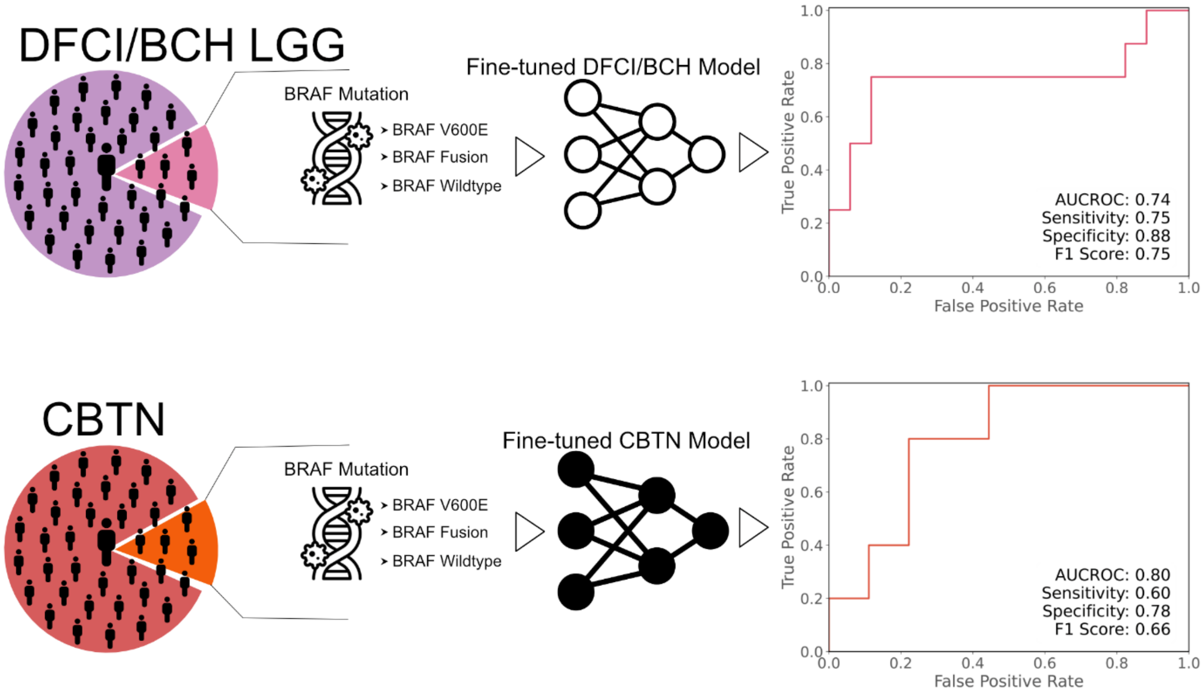

Figure S3. Comparative Analysis of BRAF Mutation Models in DFCI/BCH LGG and CBTN Datasets. The upper panel displays the distribution of BRAF mutations in a cohort of N=25 from DFCI/BCH LGG test set, with the corresponding performance of the fine-tuned BCH model depicted in a Receiver Operating Characteristic (ROC) curve (right), achieving an Area Under the ROC Curve (AUCROC) of 0.74, sensitivity of 0.75, specificity of 0.88, and an F1 score of 0.75. The lower panel shows the mutation distribution in a similar cohort of N=23 from CBTN, alongside the fine-tuned CBTN model's ROC curve, with an AUCROC of 0.80, sensitivity of 0.60, specificity of 0.78, and an F1 score of 0.66.

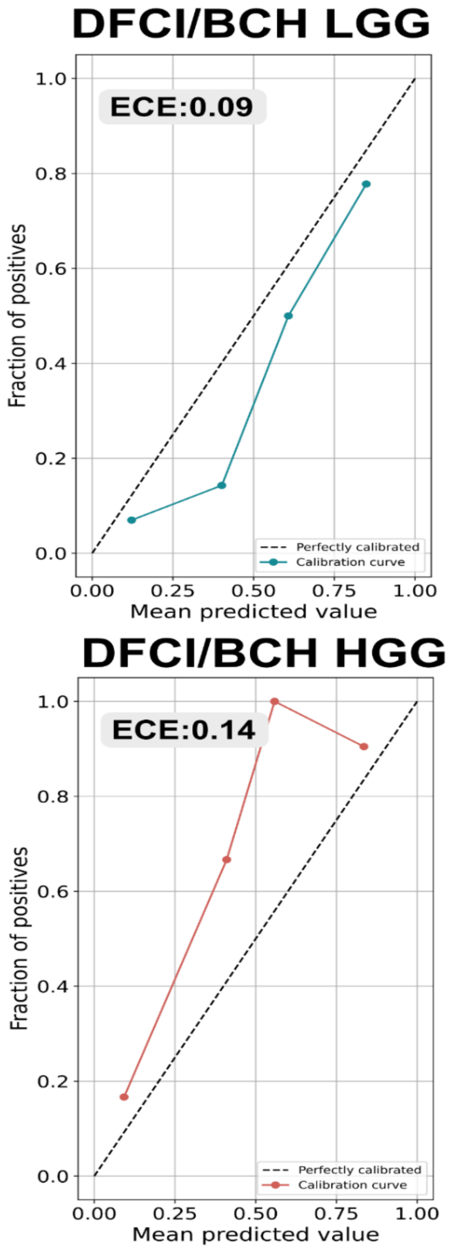

Figure S4. Calibration plots of DFCI/BCH LGG EFS Finetuned model (with temporal learning) for DFCI/BCH LGG, CBTN, DFCI/BCH HGG, RadART test sets.

#### Supplementary Tables

Table S1. Patient characteristics for DFCI/BCH LGG, CBTN, DFCI/BCH HGG, RadART cohorts

| | DFCI/BCH LGG (n=345) | CBTN (n=301) | DFCI/BCH HGG (n=34) | RadART (n=35) |
|---|---|---|---|---|
| **Age (years)** | | | | |
| Median (range) | 9 (3 – 20) | 9 (1 - 23) | 6(3-19) | 12 (5-24) |
| **Sex, n (%)** | | | | |
| Female | 170 (49.14%) | 134 (44.52%) | 13(38.24%) | 13(37.14%) |
| Male | 172 (50%) | 167 (55.48%) | 21(61.76%) | 22(62.86%) |
| Unknown | 3 (0.86%) | 0 (0%) | 0(0%) | 0(0%) |
| **Race/Ethnicity, n (%)** | | | | |
| African American/Black | 13 (3.6%) | 24 (7.97%) | | |
| Asian American/Asian | 10 (2.77%) | 5 (1.66%) | | |
| Hispanic/Latinx | 11 (3.05%) | 17 (5.65%) | | |
| Non-Hispanic Caucasian/white | 257 (71.19%) | 201 (66.78%) | | |
| other | 13 (3.60%) | 0 (0%) | | |
| Unknown | 41 (11.36%) | 54 (17.94%) | | |
| **Adjuvant Therapy (%)** | | | | |
| None | 85% | 80% | 30% | 80% |
| Chemotherapy | 5% | 9% | 0% | 9% |
| Radiation Therapy | 10% | 9% | 23% | 11% |
| Both | 1% | 2% | 47% | 0% |

Table S2 Performance comparison across datasets of different training approaches (single timepoint imaging, standard longitudinal training, temporal learning and finetuning) on both DFCI/BCH LGG and CBTN cohort.

| | AUC | Sensitivity | Specificity | F1-Score |
|---|---|---|---|---|
| **DFCI/BCH LGG (Npatient=67, Nscan=511)** | | | | |
| Single Timepoint imaging (DFCI/BCH LGG) | 0.58 [0.44-0.73] | 0.12 | 0.98 | 0.57 |
| Longitudinal Imaging (DFCI/BCH LGG) | 0.78 [0.62-0.91] | 0.64 | 0.77 | 0.67 |
| Longitudinal Imaging with temporal learning (DFCI/BCH LGG) | 0.83 [0.71-0.91] | 0.78 | 0.86 | 0.80 |
| **CBTN (Npatient=61, Nscan=204)** | | | | |
| Single Timepoint imaging (CBTN) | 0.49 [0.44-0.73] | 0.21 | 0.74 | 0.46 |
| Longitudinal Imaging (CBTN) | 0.53 [0.35-0.70] | 0.50 | 0.65 | 0.55 |
| Longitudinal Imaging with temporal learning (DFCI/BCH LGG) | 0.75 [0.58-0.90] | 0.71 | 0.60 | 0.65 |
| Longitudinal Imaging with temporal learning (CBTN) | 0.74 [0.61-0.86] | 0.78 | 0.60 | 0.67 |
| **RadART (Npatient=35, Nscan=103)** | | | | |
| Single Timepoint imaging (DFCI/BCH LGG) | 0.43 [0.25,0.67] | 0.11 | 0.98 | 0.57 |
| Longitudinal Imaging (DFCI/BCH LGG) | 0.71 [0.50,0.92] | 0.62 | 0.70 | 0.63 |
| Longitudinal Imaging with temporal learning (DFCI/BCH LGG) | 0.84 [0.68,0.95] | 0.62 | 0.85 | 0.73 |
| Single Timepoint imaging (CBTN) | 0.59 [0.48,0.70] | 0.11 | 0.79 | 0.46 |
| Longitudinal Imaging (CBTN) | 0.67 [0.56,0.78] | 0.50 | 0.67 | 0.56 |

| | | | | |
|---|---|---|---|---|
| Longitudinal Imaging with temporal learning (CBTN) | 0.71 [0.60,0.84] | 0.75 | 0.62 | 0.62 |
| **DFCI/BCH HGG (Npatient=34, Nscan=130)** | | | | |
| Single Timepoint imaging (DFCI/BCH LGG) | 0.62 [0.52,0.72] | 0.61 | 0.62 | 0.61 |
| Longitudinal Imaging (DFCI/BCH LGG) | 0.82 [0.68,0.94] | 0.88 | 0.62 | 0.75 |
| Longitudinal Imaging with temporal learning (DFCI/BCH LGG) | 0.89 [0.75,0.98] | 0.88 | 0.75 | 0.80 |
| Single Timepoint imaging (CBTN) | 0.67 [0.60,0.74] | 0.40 | 0.74 | 0.54 |
| Longitudinal Imaging (CBTN) | 0.77 [0.60,0.94] | 0.69 | 0.62 | 0.62 |
| Longitudinal Imaging with temporal learning (CBTN) | 0.81 [0.66,0.97] | 0.61 | 0.87 | 0.65 |

Table S3 Adjuvant therapy subgroup performance analysis on DFCI/BCH LGG, CBTN, DFCI/BCH HGG, RadART dataset. The adjuvant therapy subgroup consists of patients who went through adjuvant chemotherapy or radiation therapy. The best performing DFCI/BCH LGG longitudinal imaging model with temporal learning (on DFCI/BCH LGG) was tested on DFCI/BCH LGG , CBTN, RadART, and DFCI/BCH HGG test set.

| | AUC | Sensitivity | Specificity | F1-Score |
|---|---|---|---|---|
| **DFCI/BCH LGG (Npatient=67, Nscan=511)** | | | | |
| Adjuvant Therapy (16%) (Npatients=11) | 0.80 [0.60-0.92] | 0.66 | 0.87 | 0.77 |
| No Adjuvant Therapy (84%) (Npatients=56) | 0.84 [0.66-0.94] | 0.81 | 0.84 | 0.78 |
| **CBTN (Npatient=61, Nscan=204)** | | | | |
| Adjuvant Therapy (20%)(Npatients=12) | 0.57 [0.21-0.68] | 0.60 | 0.42 | 0.50 |
| No Adjuvant Therapy (80%)(Npatients=49) | 0.80 [0.65-0.94] | 0.77 | 0.62 | 0.60 |
| **RadART (Npatient=35, Nscan=103)** | | | | |
| Adjuvant Therapy (20%)(Npatients=7) | 0.75 [0.61,0.96] | 0.66 | 0.75 | 0.71 |
| No Adjuvant Therapy (80%)(Npatients=28) | 0.84 [0.63,0.97] | 0.60 | 0.82 | 0.68 |
| **DFCI/BCH HGG (Npatient=34, Nscan=130)** | | | | |
| Adjuvant Therapy (70%)(Npatients=24) | 0.91 [0.74,0.99] | 0.94 | 0.80 | 0.87 |
| No Adjuvant Therapy (30%)(Npatients=10) | 0.80 [0.62,0.94] | 0.85 | 0.66 | 0.76 |
